# FCGR2A-131H/H is under-represented amongst patients with primary immunodeficiencies

**DOI:** 10.1101/2023.10.12.23296440

**Authors:** Edward W D Flewitt, James E G Charlesworth, Smita Y Patel, Chantal E Hargreaves

## Abstract

The Fcγ receptors (FcγRs) act as modulators of the immune system and have previously been shown to play a role in immune disorders such as systemic lupus erythematosus and immune thrombocytopenic purpura. Thus far, their role in primary immunodeficiencies (PID), including common variable immunodeficiency disorders (CVID), has not been studied. In this paper we explored whether there is an association between the following single nucleotide polymorphisms (SNPs) and CVID: *FCGR2A* H131R (rs1801274), *FCGR2B* I232T (rs1050501), and *FCGR3A* F158V (rs396991). We compared the genotypes of a cohort of 83 patients with PID, including 56 with CVID, against controls. We found a significant difference between our mixed PID cohort and controls at the *FCGR2A* H131R SNP (X^2^ =7.884, p=0.019). There was not a significant difference at either of the other SNPs studied. Further, we examined the effect of FCGR SNPs on the incidence of the most common CVID complications within our cohort: anaemias, organ-specific autoimmunity, bronchiectasis, splenomegaly, granulomata, and cytopenias. We found no significant association between SNPs and the development of these complications. In summary, we have shown that there is a link between the *FCGR2A* H131R SNP and the development of a PID.

## Introduction

Fcγ receptors (FcγRs) are glycoproteins, encoded on chromosome 1q23-24, which bind the Fc portion of IgG[1, 2]. There are six human FcγRs found on various immune cells which, when ligated, modulate the immune response[2]. FcγRs vary in structure and function, and are classified according to their affinity for IgG immune complexes. The high-affinity FcγRI (CD64) binding monomeric IgG, and the low-affinity receptors, FcγRIIa (CD32a), FcγRIIb (CD32b), FcγRIIc (CD32c), FcγRIIIa (CD16a) and FcγRIIIb (CD16b)[2–5]. Engagement of the FcγRs results in cellular activation, with the exception of FcγRIIb, which is the sole inhibitory receptor. Immune cell activation is balanced by the relative engagement of the activating and inhibiting low affinity FcγRs, with immune dysregulation and autoimmune diseases such as systemic lupus erythematosus (SLE) and rheumatoid arthritis resulting from disrupting this balance[1].

There are three single nucleotide polymorphisms (SNPs) within the low-affinity receptors widely reported to affect receptor function upon IgG binding[1]. These are located in the genes *FCGR2A*, *FCGR2B*, and *FCGR3A* which encode FcγRIIa, FcγRIIb, and FcγRIIIa, respectively[6–8]. In *FCGR2A*, a histidine (H) to arginine (R) SNP (rs1801274) at amino acid 131 (NM_001136219.3(FCGR2A):c.500A>G (p.His131Arg)), in which the H variant confers higher affinity for IgG than the R variant, particularly IgG2[9]. An isoleucine (I) to threonine (T) SNP (rs1050501) is located at amino acid 232 in *FCGR2B* (NM_004001.4(FCGR2B):c.695T>C (p.Ile232Thr)), with the T variant inhibiting the receptor associating with lipid rafts in the cell membrane, reducing inhibition of B cell receptor signalling[10, 11]. Additionally, polymorphisms within the promoter region of *FCGR2B* have been linked to altered receptor expression and autoimmunity[12]. In *FCGR3A*, a phenylalanine (F) to valine (V) SNP (rs39699) at amino acid 158 (NM_001127593.1(FCGR3A):c.526T>G (p.Phe158Val)), increases the receptor’s affinity for IgG[6]. Copy number variants affecting this locus have also been described and reported to affect disease susceptibility[10, 13–16].

These SNPs are associated with an increased risk of developing autoimmune diseases. The high affinity *FCGR2A* 131H variant is associated with childhood-onset immune thrombocytopenic purpura (ITP), rheumatoid arthritis, and Kawasaki disease[17–19]. Similarly, the high affinity *FCGR3A* 158V variant is associated with both childhood- and adult-onset ITP[18, 20]. No clear pathological mechanism has been established, however variants which confer higher affinity for ligand may lead to greater immune cell activation, and thus autoimmunity. The inhibitory FcγRIIb receptor low affinity *FCGR2B* 232T variant has been associated with ITP and SLE[21–23]. In SLE the decreased inhibitory activity alters the germinal centre’s control of peripheral tolerance which is proposed to lead to the hyper-activation of the PI3K pathway, leading to the hyper-reactive features[24, 25].

Primary immunodeficiencies (PIDs) are a heterogeneous group of rare disorders characterised by poor or absent function of at least one immune system component. Common variable immunodeficiency disorders (CVID), a subtype of PID, is characterised by a marked decrease in IgG and at least one of IgM or IgA, poor antibody response to vaccination, and an onset after 2 years of age, in the absence of other causes of immunodeficiency[26, 27]. CVID has an estimated prevalence of 1 in 25,000 and is the most common symptomatic antibody deficiency disorder[28, 29]. The aetiology of CVID is not fully understood[30]. A genome-wide association study, and our whole genome sequencing study, of patients with CVID both concluded that CVID is a polygenic disease[31, 32].Patients with CVID can be broadly classed as either infections only or complex. All patients experience recurrent infections, particularly sinopulmonary infections, which often results in bronchiectasis[33]. Those with complex CVID develop disease-related complications, including autoimmunity, cytopenias, granulomatous disease and/or malignancy[34, 35]. The reduction but not absence in IgG and the presence of antibody-mediated autoimmune disease complications in some patients, suggest CVID as a disease of quality as well as quantity of antibody response. With the little antibody that is made possibly recognising self-antigen.

Given (1) that FcγR polymorphisms have been shown to modify the risk of autoimmune diseases and (2) the incidence of autoimmune complications of CVID is about 20%, including autoantibody-mediated conditions such as ITP and autoimmune haemolytic anaemia (5-8%), we hypothesise that FcγR polymorphisms may modify the disease course in CVID[36].

This study aims to:

1. Investigate the carriage of autoimmunity-associated FcγR SNPs in a cohort of patients with PID, compared to immunocompetent controls.
2. Identify whether or not there is an association with autoimmunity-associated FcγR SNPs and the development of CVID disease-related complications.

## Materials and Methods

### Subjects

DNA of 83 patients with PIDs was available for analysis. These patients were attending the Clinical Immunology outpatient department, John Radcliffe Hospital, Oxford, UK at the time of sampling. Whilst the primary focus of this study is CVID patients (n=56), other PID patients (n=27) were included as non-CVID controls. Many of these patients had clinically validated genetic diagnoses as well as disease-related complications such as cytopenias (**Table 1** and Supplementary Table 1). Diagnosis of CVID was made in accordance to the ESID guidelines[37], with patients being classified as infections only in the absence of any other complication. Patients were diagnosed with cytopenia if leukocytes or platelets were below the normal range for more than 6 months outside treatment for infection. The underlying causes of anaemia were further explored if unlikely to be PID-related. Pancytopenia was defined as all three lineages (erythrocytes, platelets, and leukocytes) below the normal range for ≥6 months. Other relevant clinical conditions were noted from patient notes. Our in-house control group consisted of 48 anonymous, healthy volunteers from Oxfordshire, UK. All individuals gave informed written consent. Studies were performed according to the Declaration of Helsinki with South Central Research Ethics Committee approval (12/SC/0044) for patient recruitment. Controls were recruited from the University of Oxford’s Nuffield Department of Medicine through the Oxford Gastrointestinal Illness Biobank (16/YH/0247).

**Table 1.** Clinical characteristics of cohort studied. †Phenotypic classification for 6 CVID patients was unable to be determined.

|  | **CVID patients (n=56)** | **non-CVID patients (n=27)** |
| --- | --- | --- |
| **Age in years (mean)** | 62 | 45 |
| **Gender (female %)** | 50 | 48.1 |
| **CVID type** |  |  |
| Infections (%)† | 28 (56) | - |
| Complex (%)† | 22 (44) | - |
| **Complications** |  |  |
| Anaemias (%) | 21 (37.5) | 4 (14.8) |
| Organ-Specific Autoimmunity (%) | 11 (19.6) | 2 (7.4) |
| Bronchiectasis (%) | 27 (48.2) | 8 (29.6) |
| Splenomegaly (%) | 23 (41.1) | 3 (11.1) |
| Granulomata (%) | 14 (25.0) | 2 (7.4) |
| Cytopenias (%) | 20 (35.7) | 5 (18.5) |

### Control cohorts

We calculated the Hardy-Weinberg equilibrium (HWE) using a Chi Squared test for all groups compared to previously published genotype frequencies and found that the patient and control group of *FCGR2A* were not within HWE (p = 0.014 and p = 0.003 respectively) (**Table 2**)[38]. To ensure a fair control group we supplemented our in-house controls with data on the same three SNPs, genotyped by the same methods, in a large previously published cohort, also shown in **Table 2**[39, 40]. All control groups were now within HWE and only the *FCGR2A* patient group was not (p = 0.014). The power, calculated from previously published population allele frequencies, equalled 1.000 for all three receptors[38].

**Table 2.**
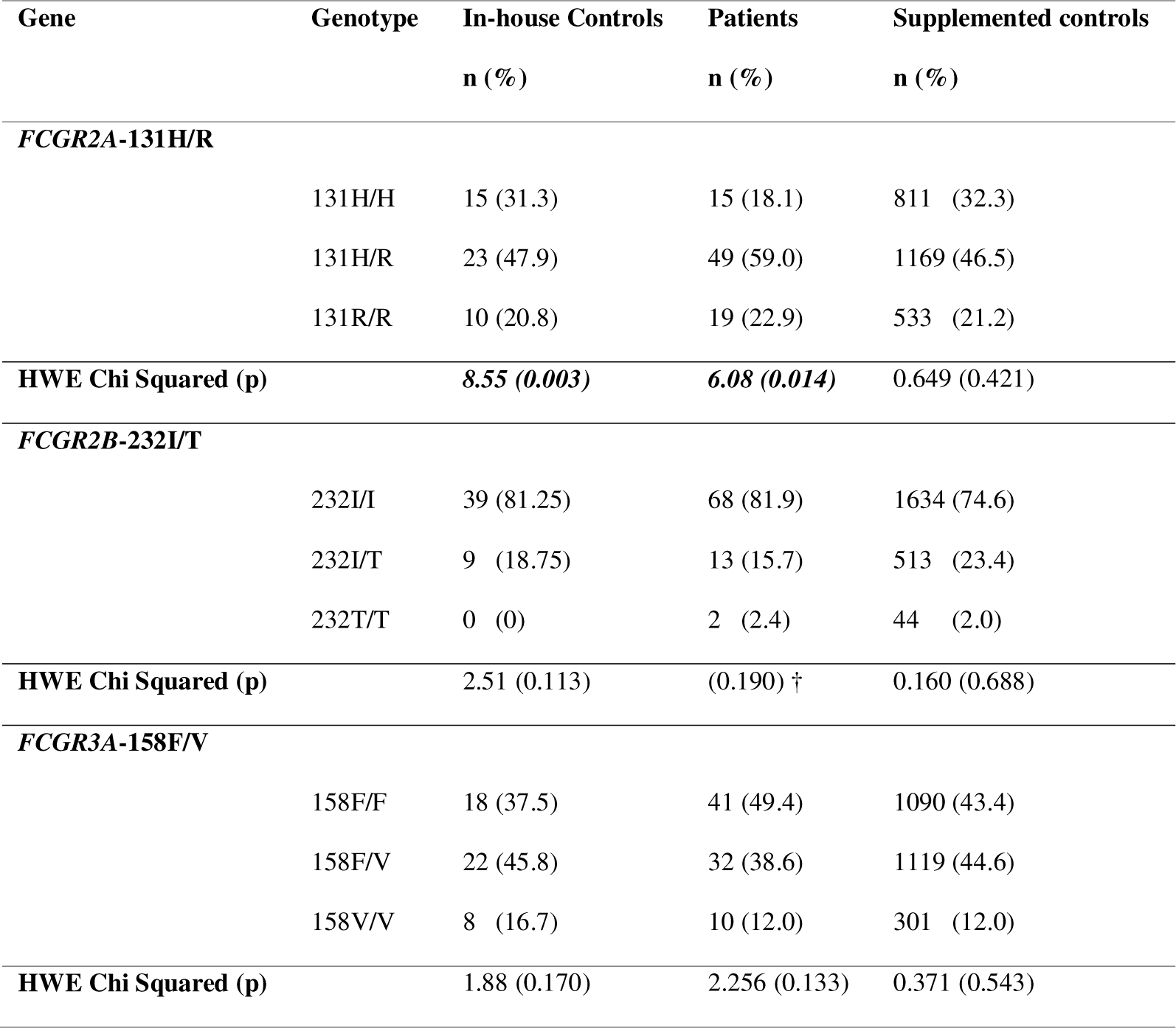
Genotype distributions and HWE calculations in patients with a PID, in-house control subjects, and total supplemented control groups. Bold italic values are statistically significant. †There were fewer than 5 individuals within a group so a HWE exact test was performed[39].

| Gene | Genotype | In-house Controls<br>n (%) | Patients<br>n (%) | Supplemented controls<br>n (%) |
| --- | --- | --- | --- | --- |
| ***FCGR2A-131H/R*** |  |  |  |  |
|  | 131H/H | 15 (31.3) | 15 (18.1) | 811 (32.3) |
|  | 131H/R | 23 (47.9) | 49 (59.0) | 1169 (46.5) |
|  | 131R/R | 10 (20.8) | 19 (22.9) | 533 (21.2) |
| **HWE Chi Squared (p)** |  | ***8.55 (0.003)*** | ***6.08 (0.014)*** | 0.649 (0.421) |
| ***FCGR2B-232I/T*** |  |  |  |  |
|  | 232I/I | 39 (81.25) | 68 (81.9) | 1634 (74.6) |
|  | 232I/T | 9 (18.75) | 13 (15.7) | 513 (23.4) |
|  | 232T/T | 0 (0) | 2 (2.4) | 44 (2.0) |
| **HWE Chi Squared (p)** |  | 2.51 (0.113) | (0.190) † | 0.160 (0.688) |
| ***FCGR3A-158F/V*** |  |  |  |  |
|  | 158F/F | 18 (37.5) | 41 (49.4) | 1090 (43.4) |
|  | 158F/V | 22 (45.8) | 32 (38.6) | 1119 (44.6) |
|  | 158V/V | 8 (16.7) | 10 (12.0) | 301 (12.0) |
| **HWE Chi Squared (p)** |  | 1.88 (0.170) | 2.256 (0.133) | 0.371 (0.543) |

### Laboratory analysis

gDNA samples were isolated from patient peripheral blood mononuclear cell samples using the Monarch Genomic DNA Purification Kit (New England Biolabs, UK) according to manufacturer’s instructions. Samples were analysed at three loci for the SNPs NM_001136219.3(FCGR2A):c.500A>G (p.His167Arg), NM_004001.4(FCGR2B):c.695T>C (p.Ile232Thr), and NM_001127593.1(FCGR3A):c.526T>G (p.Phe176Val) using a previously published TaqMan method[41]. Primers, probes, and cycling conditions are shown in Supplementary Table 2. The data was analysed using the BioRad manager software.

### Polymerase Chain Reaction (PCR) amplification

Samples were amplified using the Applied Biosystems 2720 Thermal Cycler (Applied Biosystems, CA, USA). Each reaction well contained: 1µL gDNA 5ng/µL, 12.5µL 2X GoTaq G2 Hot Start Green Master Mix (M7422, Promega, UK), 1µL forward primer 10µM (Invitrogen, UK), 1µL reverse primer 10µM (Invitrogen, UK), 1µL de-ionised H_2_O. Primer design is shown in Supplementary Table 3. Touchdown PCR was used with cycling conditions shown in Supplementary Table 4. Amplification was confirmed by gel electrophoresis.

### Gel electrophoresis

A 1X Tris-Acetate-EDTA buffer gel with 1% agarose was prepared with a 0.01% Diamond Nucleic Acid Stain (Promega, UK). PCR samples were diluted 1 in 10 with de-ionised H_2_O; these were mixed with 6X DNA Loading Dye (Thermo Scientific, UK). The samples were run for 2 hours at 110V, the Quick-Load Purple 1kb Plus Ladder (New England Biolabs, UK) was used. Samples were then imaged using the ChemiDoc UV imager (Bio-Rad Laboratories, CA, USA). A single clear band at the correct number of base pairs (*FCGR2A*: 627bp, *FCGR2B*: 751bp, *FCGR3A*: 927bp) was taken as confirmation of amplification.

### Sanger sequencing

Samples were purified using the Monarch PCR & DNA Cleanup Kit (5µg) (New England Biolabs, UK) according to manufacturer’s instructions, using 20µL of PCR product and 10µL of elution buffer. A NanoDrop 1000 (Thermo Fisher Scientific, MA, USA) full spectrum spectrophotometer was used to determine the concentration of each sample. Samples were diluted with de-ionised H_2_O to a concentration of 5-20ng/µL. The samples were Sanger sequenced at the MRC Weatherall Institute of Molecular Medicine (UK). Primer designs are shown in Supplementary Table 3. Analysis was performed using FinchTV. For seven samples, the Sanger sequence result differed from the TaqMan result. The TaqMan result was used for the *FCGR2A* and *FCGR2B* samples and the Sanger sequencing result in the *FCGR3A* samples per findings of Hargreaves et al. demonstrating superiority of the TaqMan assays for *FCGR2A* and *FCGR2B*, and the Sanger sequencing assay for *FCGR3A*[41]. For *FCGR2B* this is due to the presence of known SNPs at the Sanger sequencing probe binding sites which have been shown to disrupt the Sanger sequencing results and report an over-representation of the *FCGR2B*-232T allele compared to previously published frequencies[10, 41, 42]. One sample for *FCGR3A* was not included in the study due to Sanger sequencing returning an inconclusive result. The decision-making process is shown in Supplementary Figure 1.

### Statistics

IBM SPSS Statistics (Version 27) was used to perform Chi Squared tests; R (Version 4.0.3, pwr package 1.3-0) was used to perform power calculations. Excel (Microsoft Office) was used to calculate Hardy-Weinberg equilibrium. For all tests, p<0.05 was considered significant.

## Results

### *FCGR2A*-131H/H is under-represented amongst PID patients

Altered genotype frequencies of *FCGR2A*-R131H have been found in different patient groups as compared to controls. Specifically, an increased prevalence of the 131H allele is associated with an increased risk of diseases, including Kawasaki disease and ITP[17, 18]. We sought to establish the allele frequency amongst patients with CVID in comparison to patients with other PIDs, in-house immunocompetent controls and a previously published cohort of immunocompetent patients undergoing cancer immunotherapy[40].

For *FCGR2A*-131H/R we found a significant difference between genotype distribution in patients with a PID and controls (p=0.019), but no significant difference in allele frequencies (p=0.231) (**Table 3**). This difference was a decrease in the 131H/H genotype and a corresponding increase in the heterozygous 131H/R genotype amongst all PID patients, compared to immunocompetent controls. Subgroup analysis showed that this significant difference in genotype distribution was driven by the mixed non-CVID PID group (p=0.004) but not the CVID cohort (p=0.124) when compared to our control group (**Table 4**).

**Table 3.**
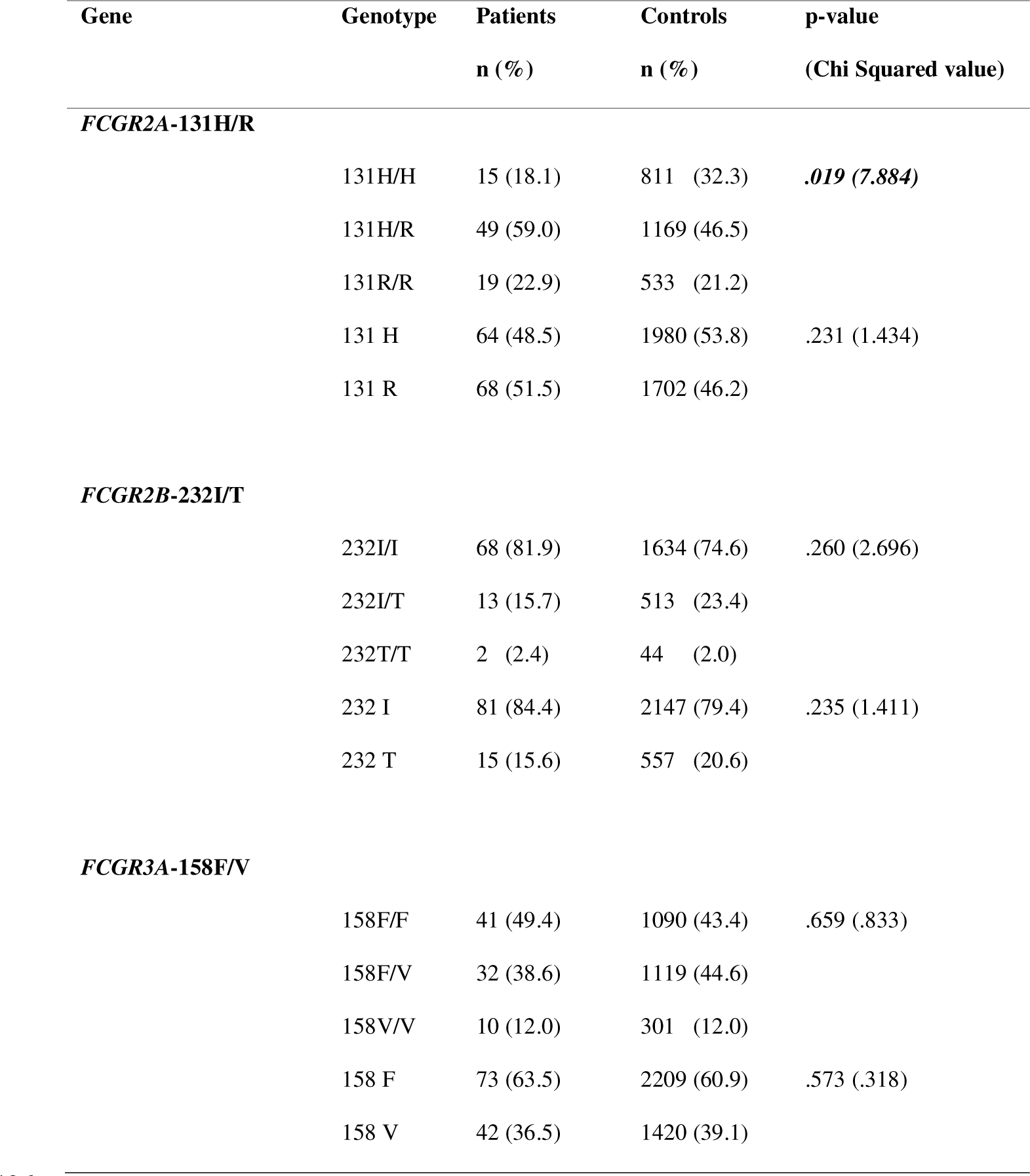
Genotype distributions and allele frequencies for patients with PIDs and controls. Bold italic values are statistically significant.

**Table 4.**
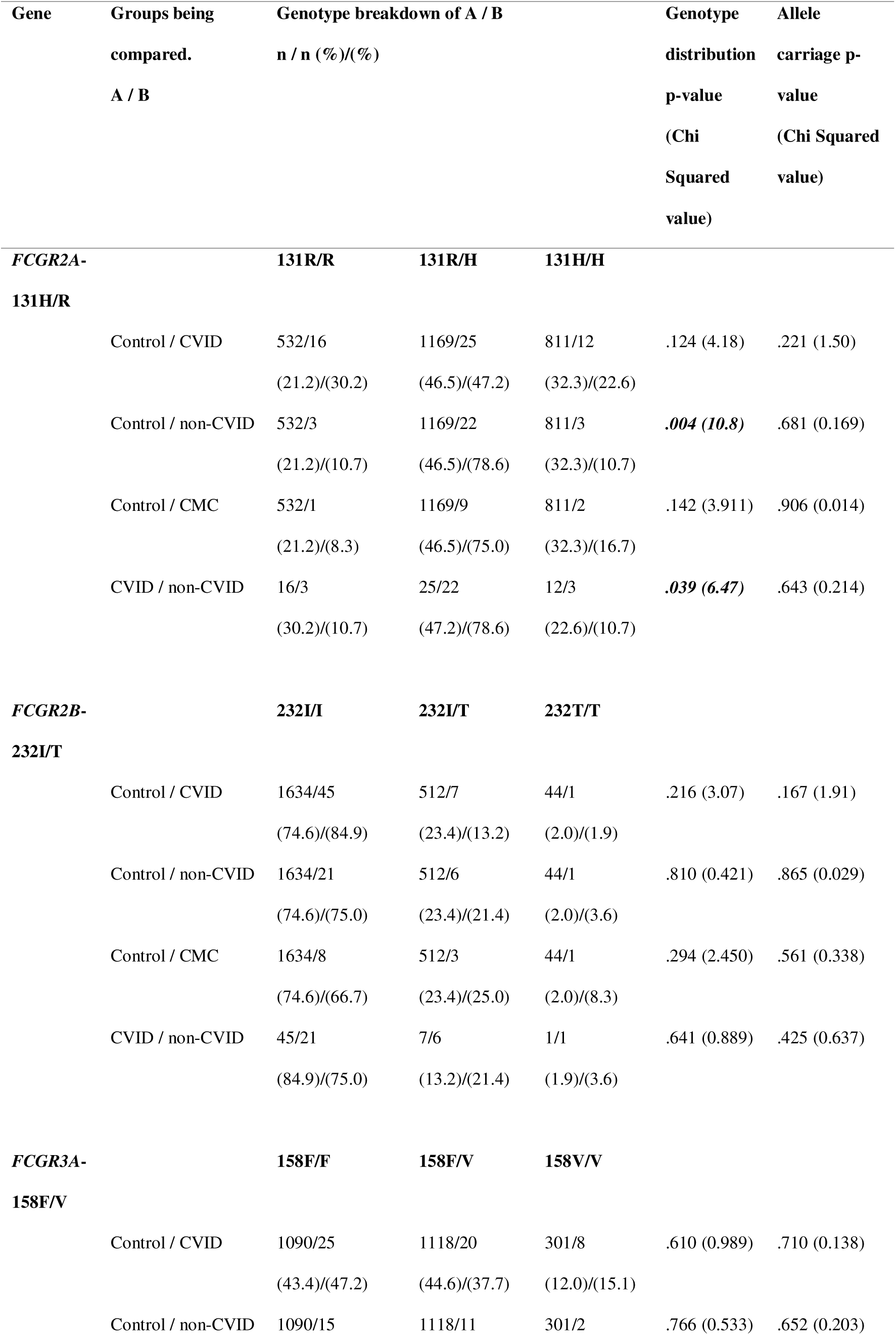
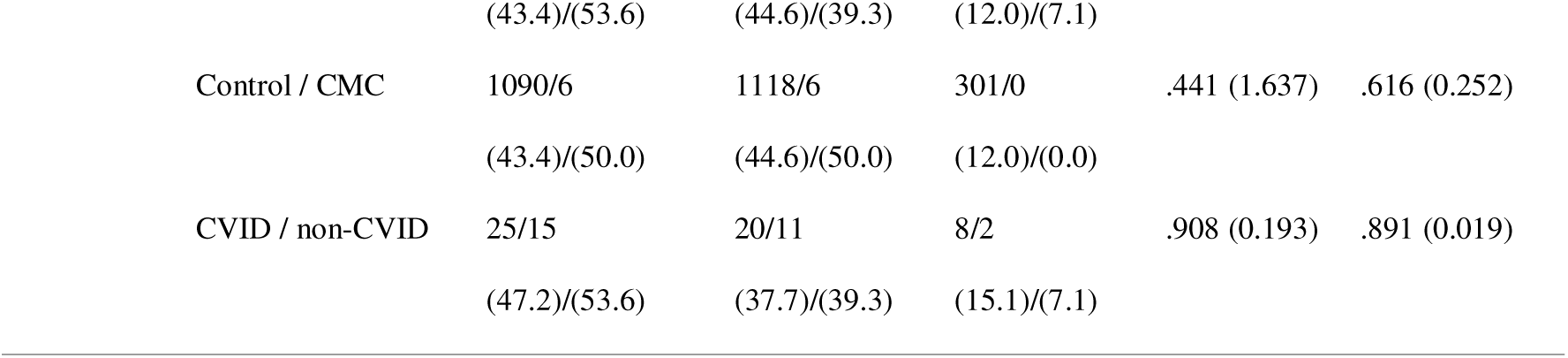
Genotype and allele carriage statistical differences between cohort groups. Bold italic values are statistically significant.

| Gene | Groups being compared.<br>A / B | Genotype breakdown of A / B<br>n / n (%) / (%) |  |  | Genotype distribution<br>p-value<br>(Chi Squared value) | Allele carriage p-value<br>(Chi Squared value) |
| --- | --- | --- | --- | --- | --- | --- |
| **FCGR2A-131H/R** |  | **131R/R** | **131R/H** | **131H/H** |  |  |
|  | Control / CVID | 532/16<br>(21.2)/(30.2) | 1169/25<br>(46.5)/(47.2) | 811/12<br>(32.3)/(22.6) | .124 (4.18) | .221 (1.50) |
|  | Control / non-CVID | 532/3<br>(21.2)/(10.7) | 1169/22<br>(46.5)/(78.6) | 811/3<br>(32.3)/(10.7) | **.004 (10.8)** | .681 (0.169) |
|  | Control / CMC | 532/1<br>(21.2)/(8.3) | 1169/9<br>(46.5)/(75.0) | 811/2<br>(32.3)/(16.7) | .142 (3.911) | .906 (0.014) |
|  | CVID / non-CVID | 16/3<br>(30.2)/(10.7) | 25/22<br>(47.2)/(78.6) | 12/3<br>(22.6)/(10.7) | **.039 (6.47)** | .643 (0.214) |
| **FCGR2B-232I/T** |  | **232I/I** | **232I/T** | **232T/T** |  |  |
|  | Control / CVID | 1634/45<br>(74.6)/(84.9) | 512/7<br>(23.4)/(13.2) | 44/1<br>(2.0)/(1.9) | .216 (3.07) | .167 (1.91) |
|  | Control / non-CVID | 1634/21<br>(74.6)/(75.0) | 512/6<br>(23.4)/(21.4) | 44/1<br>(2.0)/(3.6) | .810 (0.421) | .865 (0.029) |
|  | Control / CMC | 1634/8<br>(74.6)/(66.7) | 512/3<br>(23.4)/(25.0) | 44/1<br>(2.0)/(8.3) | .294 (2.450) | .561 (0.338) |
|  | CVID / non-CVID | 45/21<br>(84.9)/(75.0) | 7/6<br>(13.2)/(21.4) | 1/1<br>(1.9)/(3.6) | .641 (0.889) | .425 (0.637) |
| **FCGR3A-158F/V** |  | **158F/F** | **158F/V** | **158V/V** |  |  |
|  | Control / CVID | 1090/25<br>(43.4)/(47.2) | 1118/20<br>(44.6)/(37.7) | 301/8<br>(12.0)/(15.1) | .610 (0.989) | .710 (0.138) |
|  | Control / non-CVID | 1090/15 | 1118/11 | 301/2 | .766 (0.533) | .652 (0.203) |

**Table 4. Genotype and allele carriage statistical differences between cohort groups.** Bold italic values are statistically 430 significant.
|  |  |  |  |  |  |
| --- | --- | --- | --- | --- | --- |
|  | (43.4)/(53.6) | (44.6)/(39.3) | (12.0)/(7.1) |  |  |
| Control / CMC | 1090/6 | 1118/6 | 301/0 | .441 (1.637) | .616 (0.252) |
|  | (43.4)/(50.0) | (44.6)/(50.0) | (12.0)/(0.0) |  |  |
| CVID / non-CVID | 25/15 | 20/11 | 8/2 | .908 (0.193) | .891 (0.019) |
|  | (47.2)/(53.6) | (37.7)/(39.3) | (15.1)/(7.1) |  |  |

There was a statistically significant difference between the genotype distribution of the mixed non-CVID PID group and the CVID group (p=0.039), with the 131R/R genotype making up 30.2% of CVID patients compared to just 10.7% of non-CVID PID patients (**Table 4**). There was no significant difference in allele frequencies upon any subgroup analysis (**Table 4**).

Further subgroup analysis of genotype and allele distribution based upon clinical phenotypes found no significant differences (**Table 5**).

**Table 5.**
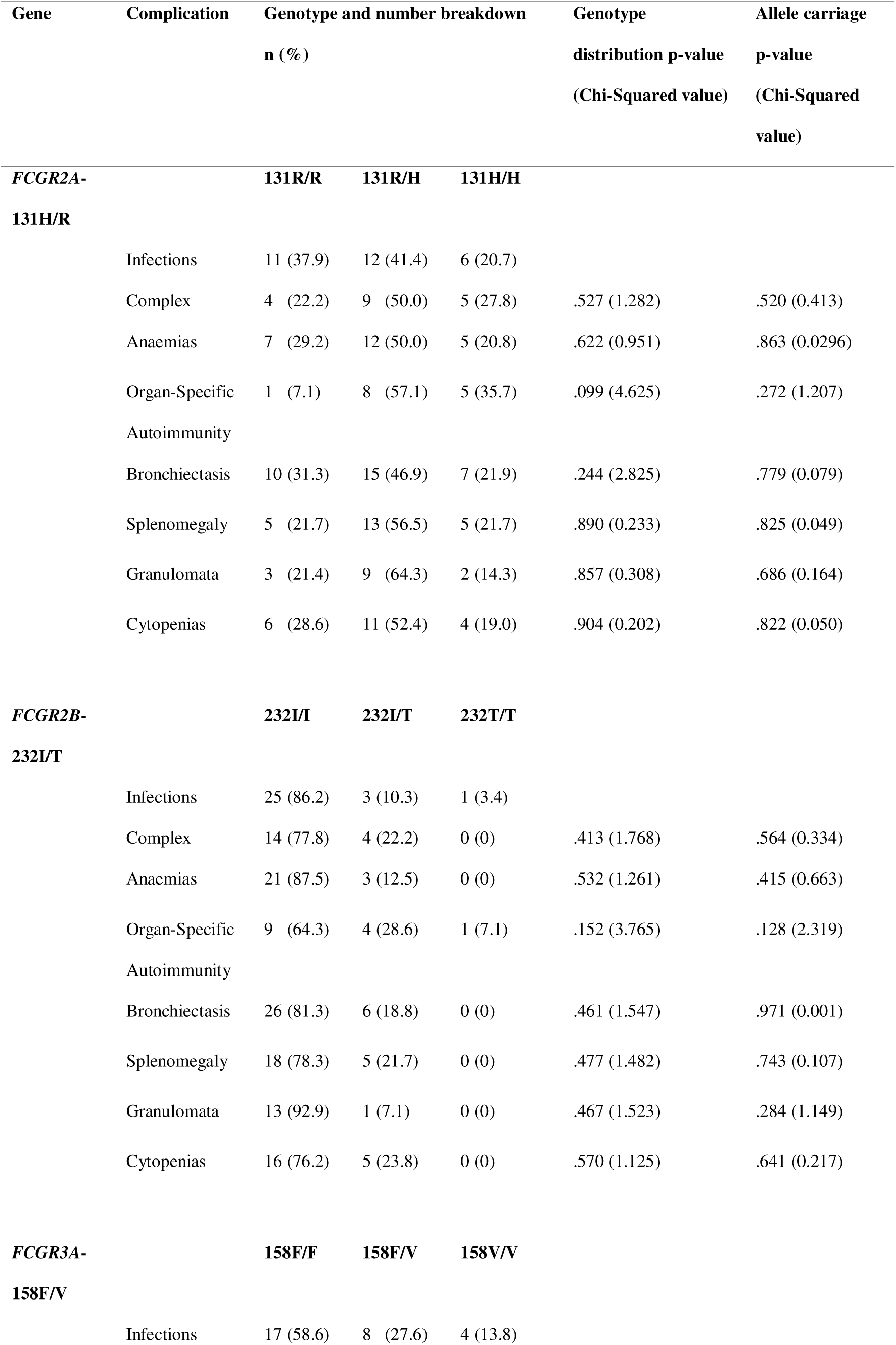
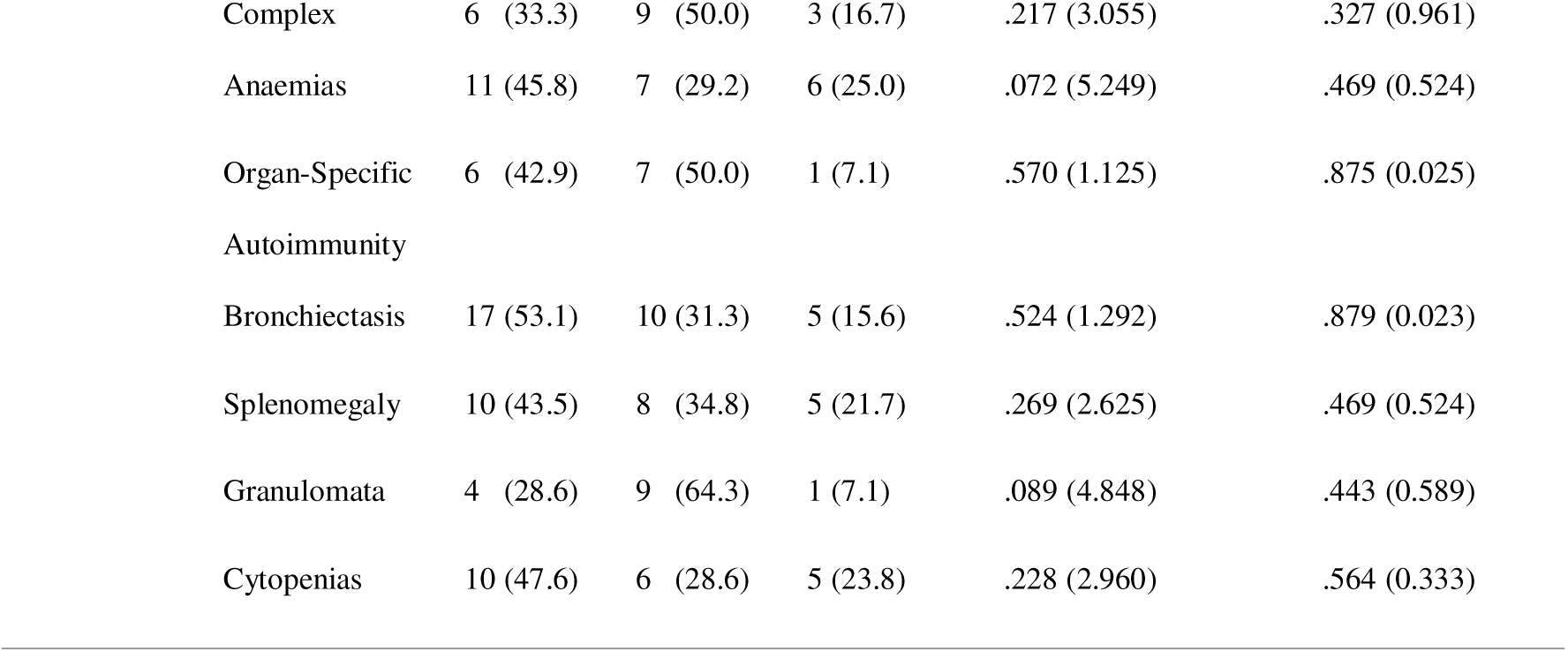
Analysis of complications development, broken down by SNP, within the PID cohort.

### In PID patients there was no significant difference in *FCGR2B*-232I/T and *FCGR3A*-158F/V compared to controls

Altered genotype frequencies of *FCGR2B*-232I/T and *FCGR3A*-158F/V have been found in patient groups compared with controls. The low affinity 232T variant has been associated with an increased risk of ITP and SLE[21–23]. Whilst the high affinity 158V variant has been shown to be associated with both childhood- and adult-onset ITP[18, 20]. We sought to establish the allele frequency amongst patients with CVID in comparison to patients with other PIDs, in-house immunocompetent controls and a previously published cohort of immunocompetent patients undergoing cancer immunotherapy[40].

For both *FCGR2B*-232I/T and *FCGR3A*-158F/V we found no significant difference in genotype distribution (p=0.260, p=0.659) or allele frequencies (p=0.235, p=0.573) compared to controls (**Table 3**). Subgroup analysis breaking down the PID population into CVID, non-CVID and CMC groups also yielded no significance in either genotype distribution or allele carriage (**Table 4**) and neither did analysis based upon clinical phenotype (**Table 5**).

## Discussion

The clinical significance of low affinity FcγR in disease has been investigated by various groups over the last 20 years[43]. Previous studies in Kawasaki disease and ITP found that the *FCGR2A*-131R/R genotype is linked with increased disease risk[17, 18]. The mechanisms behind this association have not been explored. However, due to the differing affinities between these receptors it leads to variation in the clearing of immune complexes, antigen presentation, and antibody mediated phagocytosis all of which play a role in autoimmune diseases[20]. It is possible that the greater proportion of the high affinity allele could result in an increased clearance of IgG sensitized cells. Thus, leading to an overactivation of the immune system which could cause autoimmune attacks which impair aspects of the immune system, causing a PID to develop.

We found no association between *FCGR2A* polymorphisms and CVID incidence rates (p=0.124). Although our data did support an association between *FCGR2A* polymorphism and the presence of any PID compared to control (p=0.004). This association was a decrease in the proportion of the high affinity 131H/H receptor within the patient group compared to controls. Whilst this has not been previously explored, the finding was interesting given the opposite previous findings in ITP and Kawasaki’s disease[17, 18]. Due to the relatively small non-CVID cohort we were unable to further explore this association, however, this result does imply that the mechanism driving low platelet counts could differ between CVID and non-CVID ITP. Also, given the majority of non-CVID PIDs within our patient cohort had CMC (Supplementary Table 1) there is a possibility that this polymorphism is linked to inhibition of the Th17 and Th1 immune cell pathways which have previously been implicated in CMC progression[44]. To prove such a link would require further research into the area.

We found no significant differences between the CVID and PID cohorts and controls in both *FCGR2B* (p=0.260) and *FCGR3A* (p=0.659). This may be due to *FCGR2B* and *FCGR3A* not contributing to pathology in this cohort, or our it could be that our cohort is simply too small to detect an association driven by these variants.

We supplemented our controls with data collected from Strefford et al[40]. The patients involved in this study were being treated with either obinutuzumab- or rituximab-chemotherapy for untreated follicular lymphoma and diffuse large B-cell lymphoma in the GALLIUM and GOYA trials[45, 46]. While the cohort are not healthy individuals; the median age of patients in these studies (GALLIUM=59 years, GOYA=62 years) was significantly higher than the typical age of diagnoses for CVID (20-40 years) or other PIDs (median 9.5 years)[47, 48]. It is therefore unlikely that undiagnosed PID cases could be within the reference control cohort. FcγR genetics have never been demonstrated to predispose to cancer, and indeed the genotype frequencies were within expected frequencies for this cohort, suggesting there was no selection pressure as demonstrated amongst autoimmune cohorts. Furthermore, neither cancer is directly linked to the presence of a PID and therefore for our purposes these patients form a suitable control group.

When comparing the Sanger sequence traces to the TaqMan data there were occasional discrepancies between the two readouts. For both *FCGR2A* and *FCGR2B* we prioritised the readout from the TaqMan assays as this has been previously validated[41]. For *FCGR3A,* there is copy number variation which can complicate distinguishing FFV and FVV genotypes from FV[13]. Sanger sequencing is less affected, providing the sequence from each allele can be detected, and thus this was prioritised in cases of discrepancy[41].

### Study limitations

The small sample size was a limitation of this study, especially within the non-CVID patient cohort, as this limited the analysis which could be performed on the statistically significant difference found at the *FCGR2A* SNP. The number of different diseases present within the non-CVID patients (Supplementary Table 1) is also potentially problematic given that the different genetic make-up of each disease could affect the FcγRs differently.

A further limitation of the study is the discrepancies present between our TaqMan and Sanger sequencing results. We are not sure why we had a higher rate of discrepancy than researchers who have previously used the same methodology[40]. However, to mitigate the effect of these discrepancies future work could involve the Sanger sequencing of a larger percentage of samples. Given that in both *FCGR2A* and *FCGR3A* it was the same patient sample which was discordant this could be due to a case of individual genetic variation, rather than a systemic issue. Finally, a repeat of the genotyping using a different set of primers could lead to a set of concordant results. For instance, a nested PCR protocol could lead to increased consistency in results due to its potential for higher sensitivity and specificity[49].

### Further work

This work could be expanded upon by increasing patient numbers, particularly the number of non-CVID patients, and exploring a wider range of PID related complications. Doing so could allow for a greater insight to the rare *FCGR2B*-232T/T genotype, as well as the rarer complications observed. However, given that PIDs are rare diseases, this would certainly require collaboration with a group(s) from a different geographical area as we were able to access samples from the majority of consenting PID patients who have lived within the John Radcliffe catchment area in the last decade. Alternatively, given the increasing number of PID patients who have undergone whole genome sequencing this could be an avenue for further analysis of the FcγR SNPs mentioned[32].

Further, we did not consider the remaining two low affinity FcγRs: *FCGR2C* and *FCGR3B*. Both genes contain disease-associated SNPs and copy number variations[50]. These differences have been shown in previous studies to play a role in immune diseases such as SLE and ITP and thus may potentially play a role in CVID development given their expression on natural killer cells and neutrophils, something that could be explored in future research[3, 51, 52].

### Conclusions

In summary, our results show that there is no significant genetic role of SNPs on the *FCGR2A*, *FCGR2B*, and *FCGR3A* genes in CVID. We have shown that these SNPs both do not affect the risk of developing CVID and are also not implicated in an increased risk of developing any of the most common complications. We found evidence suggesting that *FCGR2A* may play a role in the development of PIDs, but further studies of these FcγR SNPs are needed to confirm this finding.

**Figure 1.**
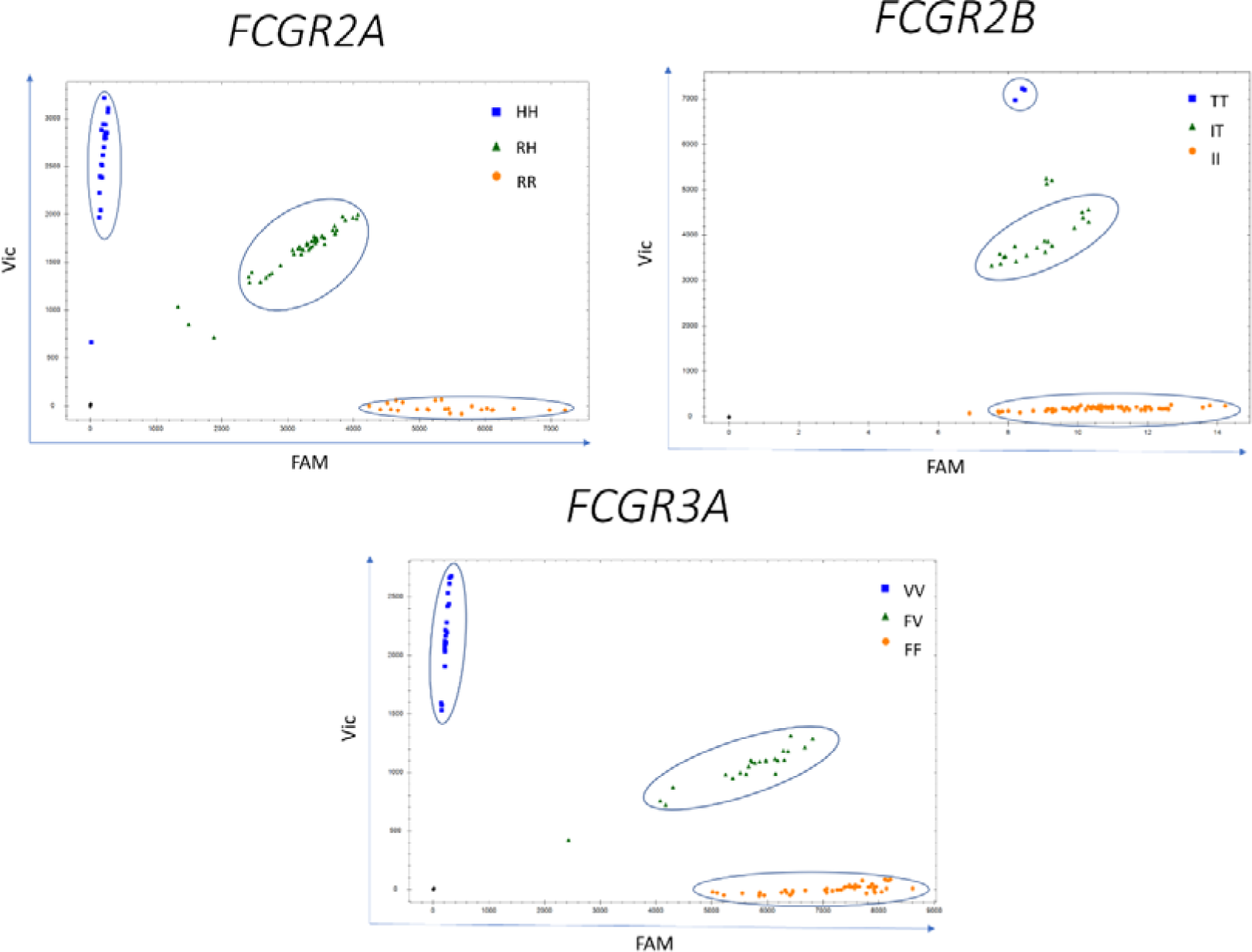
TaqMan genotyping plots with gating strategy shown. gDNA samples were genotyped in triplicate for genes *FCGR2A*, *FCGR2B,* and *FCGR3A*. The reference alleles for each gene are shown in orange circles, the alternate allele in blue squares and heterozygotes in green triangles and non-template controls as black diamonds. Gates were manually drawn to genotype samples. Samples which were not easily gated were confirmed by Sanger sequencing.

## Data Availability

The data that support the findings of this study are available from the corresponding author upon reasonable request.

## Abbreviations

CMC: chronic mucocutaneous candidiasis
CVID: common variable immunodeficiency disorder
F: phenylalanine
FCGR: Fc gamma receptor
H: histidine
HWE: Hardy-Weinberg equilibrium
I: isoleucine
ITP: immune thrombocytopenia purpura
PCR: polymerase chain reaction
PID: primary immunodeficiencies
R: arginine
SLE: systemic lupus erythematosus
SNP: single nucleotide polymorphism
T: threonine
V: valine

## Ethics Statement

Control and patient blood samples and gDNA were collected and stored with written informed consent.

## Author Contributions

EWDF performed experiments, analysed data and wrote the manuscript; JEGC collected clinical information; CEH and SYP conceived and designed the study, supervised the study and wrote the manuscript. All authors critically reviewed and approved the final version of the manuscript.

## Acknowledgements

The authors would like to acknowledge Alex Adams, Translational Gastroenterology Unit, Nuffield Department of Medicine for statistical advice.

