## Supplementary Tables 1-4 for "FCGR2A-131H/H is under-represented amongst patients with primary immunodeficiencies"

| Disease(s) | Number of patients |
| --- | --- |
| Common Variable Immunodeficiency Disorders | 56 |
| Chronic Mucocutaneous Candidiasis | 13 |
| C8 Deficiency | 1 |
| DOCK8 Deficiency | 1 |
| Janus Kinase 3 Deficiency | 1 |
| NFKB1/PIK3R1 deficiency | 1 |
| Primary Antibody Deficiency | 3 |
| SAMD9L mutation | 1 |
| Secondary Immunodeficiency to non-Hodgkin’s Lymphoma | 1 |
| Specific polysaccharide antibody deficiency syndrome | 2 |
| X-linked inhibitor of apoptosis protein deficiency | 1 |
| X-linked agammaglobulinemia | 2 |

**Supplementary Table 1.** Breakdown of diseases within the patient cohort.

| Temp. (**°**C) | Time (m:s) | Cycles |
| --- | --- | --- |
| 95 | 10:00 | 1 |
| 95 | 0:15 | 40 |
| 60 | 01:00 |  |
| Plate read | - |  |

**Supplementary Table 2.** qPCR cycling conditions.

| **Gene/SNP** | **Primer** | **Primer Sequence** |
| --- | --- | --- |
| ***FCGR2A*** **R131H**  rs1801274 | Forward PCR and sequencing primer | CATATATTGCCTATAAGAGAATGCT |
|  | Reverse PCR and sequencing primer | CCTGACTACCTATTACCTGGGA |
| ***FCGR2B* I232T**  rs1050501 | Custom forward TaqMan primer | GATGGGGATCATTGTGGCTGT |
|  | Custom reverse TaqMan primer | AGGCCACTACAGCAGCAACAAT |
|  | Custom TaqMan 232I probe | ACTGGGATTGCTGTAGC |
|  | Custom TaqMan 232T probe | ACTGGGACTGCTGTAGC |
|  | Forward PCR and sequencing primer | CTGCCTGCTCACAAATGTA |
|  | Reverse PCR and sequencing primer | CACTGCTCTCCCCAAGAC |
| ***FCGR3A*** **F158V**  rs396991 | Forward PCR primer | CCTCTAATAGGGCAATTCATCATT |
|  | Reverse PCR and sequencing primer | AGATGTGGCTTCTGCTCCTG |
|  | Forward sequencing primer | TGCTCTGCATAAGGTCACATATT |

**Supplementary Table** **3.** PCR primer designs, TaqMan probe designs, and Sanger sequencing primer designs.

| Temp. (**°**C) | Time (m:s) | Cycles |
| --- | --- | --- |
| 95 | 10:00 | 1 |
| 95 | 0:30 | 8 |
| 65 – 1°C drop per cycle | 0:45 |  |
| 72 | 1:30 |  |
| 95 | 0:30 | 27 |
| 57 | 0:45 |  |
| 72 | 1:30 |  |
| 72 | 5:00 | 1 |
| 4 | Hold | |

**Supplementary Table 4.** PCR cycling conditions.
