## Supplementary Figure 1 for "FCGR2A-131H/H is under-represented amongst patients with primary immunodeficiencies"

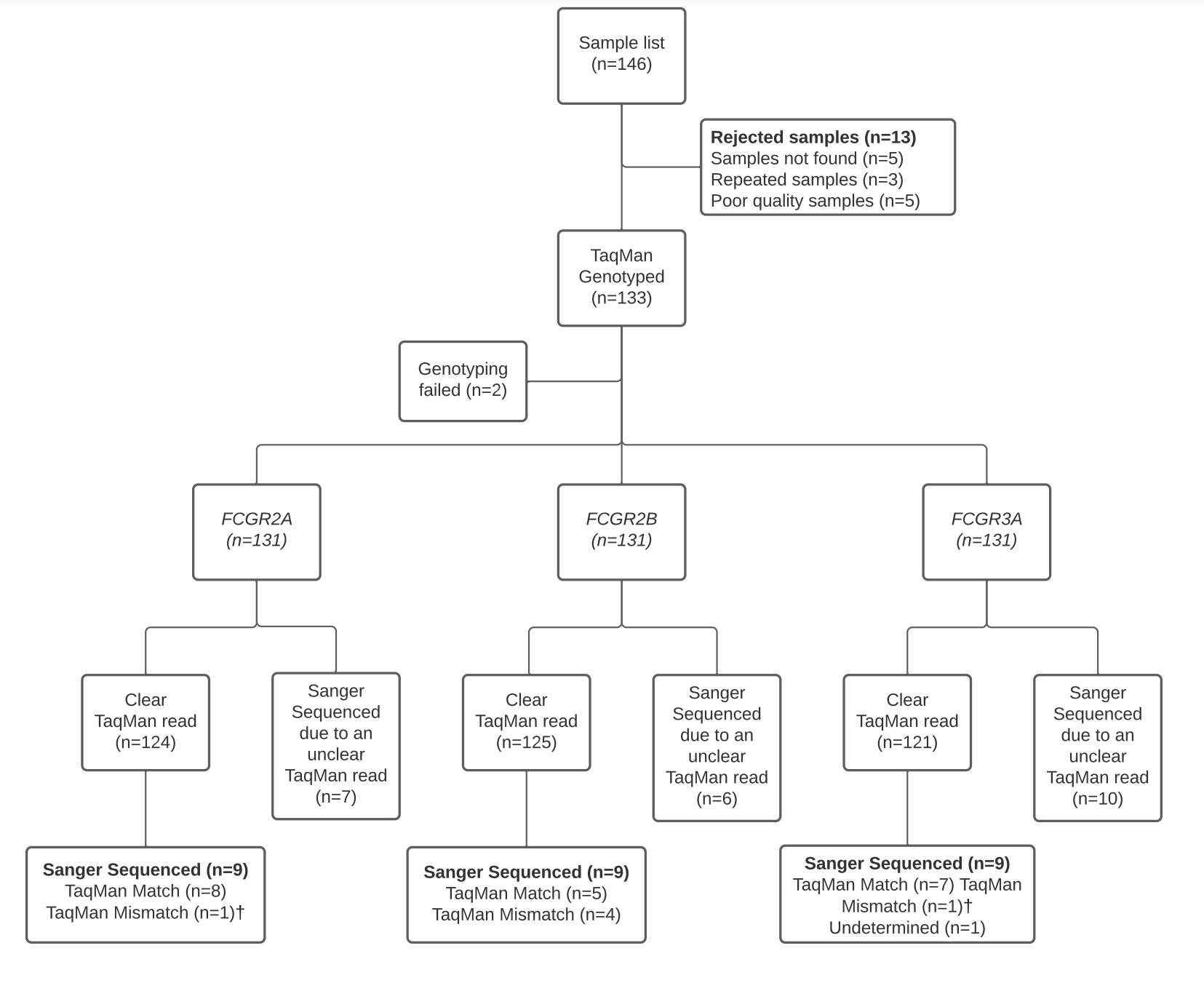

**Supplementary Figure 1. A flowchart showing a summary of the study process followed.** †The same sample had a TaqMan Mismatch in both FCGR2A and FCGR3A.
